# Trends in gestational age at birth in the city of São Paulo, Brazil between 2012 and 2019

**DOI:** 10.1101/2025.02.02.25321548

**Authors:** Margarida Maria Tenório de Azevedo Lira, Marina de Freitas, Eliana de Aquino Bonilha, Célia Maria Castex Aly, Patrícia Carla dos Santos, Denise Yoshie Niy, Carmen Simone Grilo Diniz

## Abstract

Studies have shown that excessive obstetric interventions such as induced labor and caesarean sections have contributed to the shortening of the length of gestation, leading to a left shift in gestational age (GA) at birth. The aim of this study was to analyze trends in GA and the contribution of associated factors to changes in São Paulo city, Brazil during the period 2012-2019. We conducted an ecological time-series study of births in São Paulo using data from Brazil’s national live births information system (SINASC). We calculated the average annual percent change in the proportion of births by GA between 2012 and 2019 and between the first and second four-year periods of the time series by applying log transformation to the percentages, followed by Prais-Winsten regression. A total of 1,525,759 live births were analyzed. From 2015, there was an increase in the proportion of live births between 39 and 40 weeks from 2015 and a fall in the proportion of early term (37-38 weeks) and preterm (< 37 weeks) births throughout the study period. The average annual increase in the proportion of births at 39 and 40 weeks was 7.9% and 5.7%, respectively, while the proportion of births at other gestational ages showed a statistically significant reduction over the study period. These reductions were more pronounced in the first four-year period (2012-2015). The same trend was observed when the data were analyzed by type of delivery, type of service (public or private), maternal age, and maternal education level. The findings show that there was a right shift in the GA curve during the study period and a reduction in the proportion of preterm and early term births. These changes were more pronounced in births that occurred in private hospitals. These changes reflect public policies implemented to reduce obstetric interventions such as induced labor and caesarean section before labor, especially before 39 weeks of gestation.

## Introduction

Length of gestation is one of the leading predictors of newborn health outcomes. Gestational age (GA) can be estimated using different methods and is generally measured in completed weeks. Until recently, GA was treated as a binary question, with newborns being considered either preterm (< 37 weeks) or term (37 0/7 to 41 6/7 weeks), justifying interventions such as labor induction or caesarean section from 37 weeks, when fetal development may not be complete. Studies have shown that the term period should not be treated uniformly, as early term infants (37-38 weeks) tend to have a higher risk of morbidity and mortality than those born between 39 and 41 weeks, often showing similar outcomes to late preterm infants [1–7]. In 2013, the American College of Obstetricians and Gynecologists [8] proposed a different classification of GA, dividing the term period into three categories: early term (37-38 weeks), full term (39-40 weeks), and late term (41 weeks).

Brazil’s national live births information system (SINASC) used to record GA as a categorical variable in weekly intervals up to 2010, until the Certificate of Live Birth (CLB) was modified to record this information as a continuous variable. This change enabled a more comprehensive analysis of the distribution of GA. In the city of São Paulo, the new CLB began to be used in 2011, but full implementation across all health facilities only occurred in 2012.

The average physiological length of pregnancy is 280 days, or 40 weeks [9,10]. However, interventions such as scheduled labor induction or caesarean section can shorten the length of gestation [7,11,12]. In Brazil, caesarean section rates have been far higher than the World Health Organization recommended rate of 15% for decades, reaching 57% in the general population [13,14].

In a study investigating the contribution of private sector deliveries in the city of São Paulo to reductions in length of gestation, Diniz et al. [5] found a one-week left shift in GA in infants born by caesarean section through the private health system; the same result was reported by Raspantini et al [15]. A study in Australia [7] investigating trends in the distribution of GA and the contribution of planned births (induced labor and elective caesarean section) to changes also observed a left shift in GA at birth and the findings suggest a changing pattern towards fewer births commencing spontaneously and increasing planned births.

The aim of this study was to analyze trends in GA at birth and the contribution of associated factors in the city of São Paulo during the period 2012-2019.

## Materials and methods

### Type of study and data source

We conducted an ecological time-series study of births that occurred in the city of São Paulo (SP) in the period 2012-2019 using data from the SINASC. Data were accessed for research purposes in 22/03/2022.

### Study population

We studied liveborns delivered in public and private hospitals at 22 to 45 weeks of gestation weighing ≥ 500 grams born to mothers aged between 10 and 49 years. Births without information on type of delivery, GA, and type of pregnancy were excluded (Table 1).

**Table 1.**
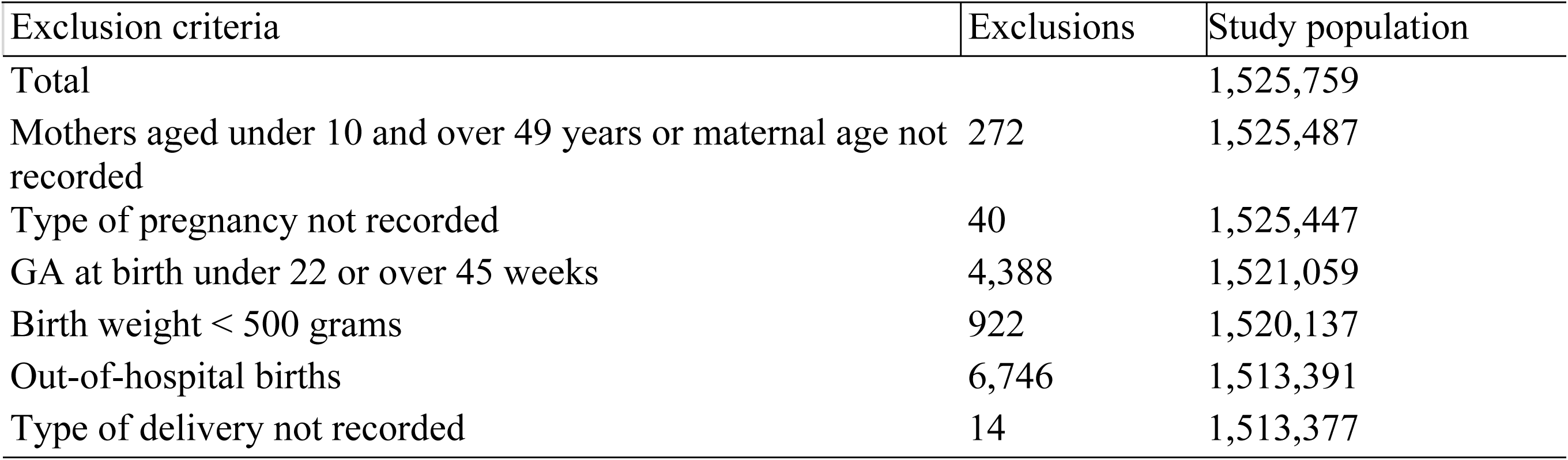
Study population and exclusions. City of São Paulo, Brazil, 2012-2019.

| Exclusion criteria | Exclusions | Study population |
| --- | --- | --- |
| Total |  | 1,525,759 |
| Mothers aged under 10 and over 49 years or maternal age not recorded | 272 | 1,525,487 |
| Type of pregnancy not recorded | 40 | 1,525,447 |
| GA at birth under 22 or over 45 weeks | 4,388 | 1,521,059 |
| Birth weight $< 500$ grams | 922 | 1,520,137 |
| Out-of-hospital births | 6,746 | 1,513,391 |
| Type of delivery not recorded | 14 | 1,513,377 |

A total of 1,525,759 live births were recorded in SP between 2012 and 2019. Of these, 12,382 (0.8%) were excluded because they did not meet the inclusion criteria (mothers aged under 10 or over 49 years or maternal age not recorded, 272; type of pregnancy not recorded, 40; GA at birth under 22 or over 45 weeks, 4,388; birth weight < 500 grams, 922; out-of-hospital births, 6,746; and type of delivery not recorded, 14), resulting in a final sample of 1,513,377 (99.2%) (Table 1).

The results of the exploratory analysis showed that the GA curves for all live births and singleton births overlapped. These types of pregnancy were therefore analyzed together for the purposes of the present study (Fig 1).

**Figure 1.**
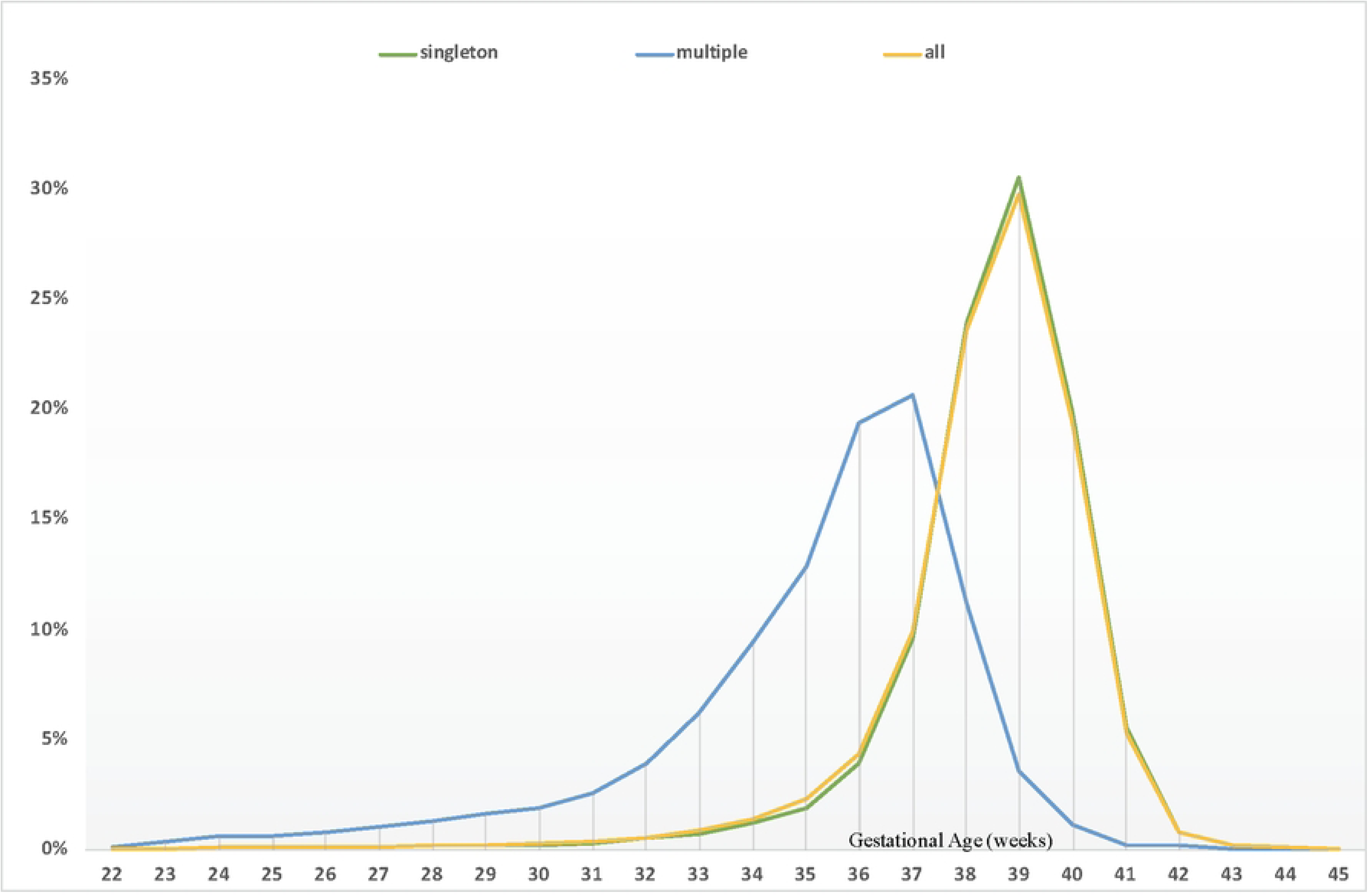
Live births by type of pregnancy. City of São Paulo, Brazil, 2012-2019.

### Statistical analysis

First we performed a descriptive statistical analysis of the data, followed by an analysis of trends in GA at birth by calculating the average annual percent change in the proportion of live births over the period 2012-2019. The results showed an increase in the proportion of births between 39 and 40 weeks from 2015. Differences in percent change rates were particularly pronounced between private and public sector births and type of delivery (data not shown). We therefore opted to compare the first and last four-year periods (2012-2015 and 2016-2019, respectively) (Fig 2).

**Figure 2.**
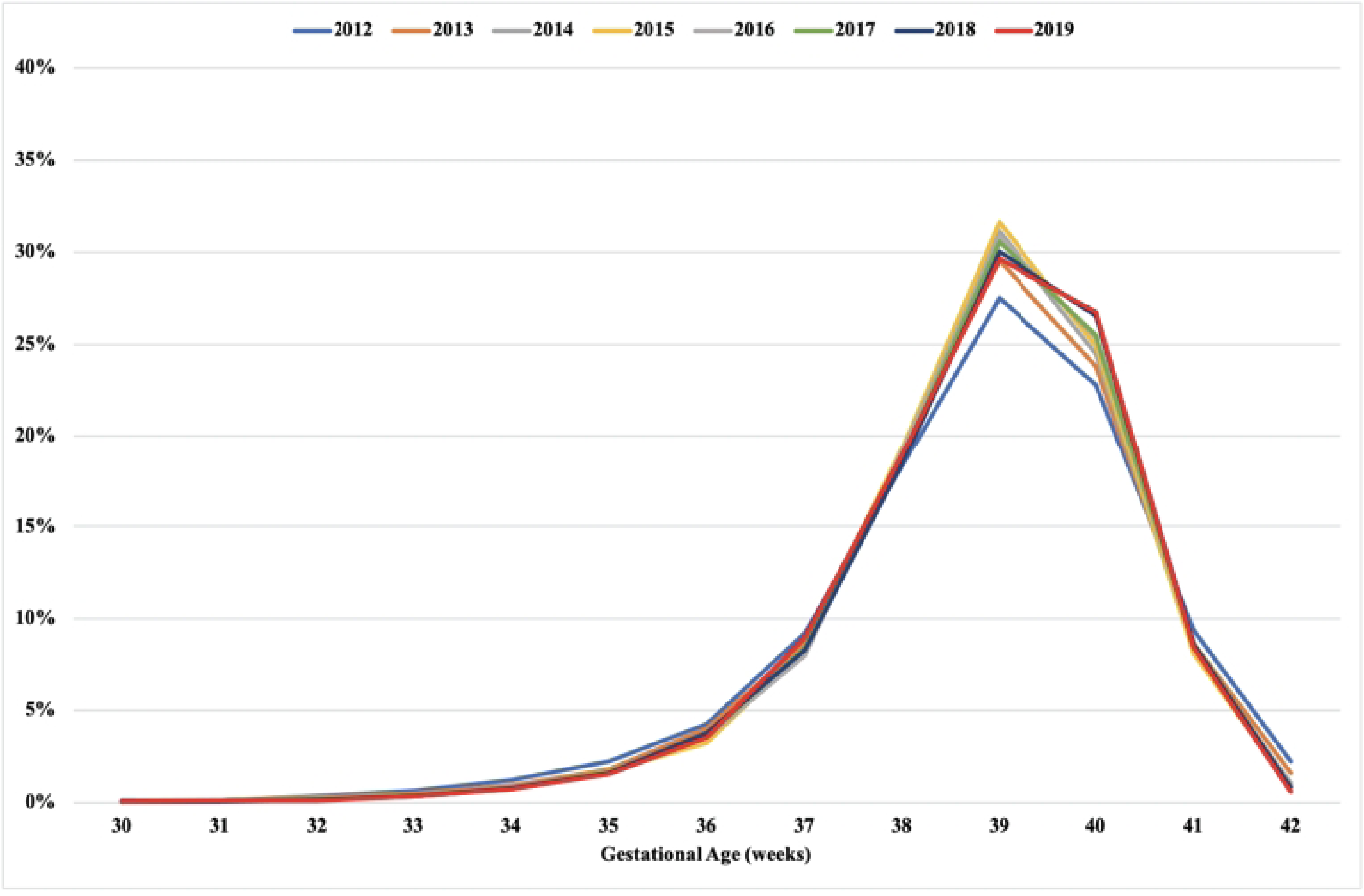
Distribution of live births by gestational age. City of São Paulo, Brazil, 2012-2019.

Percent change rate was calculated by applying log transformation to the percentages, followed by Prais-Winsten regression [16] to estimate annual percent change (**β**1) in births by GA. Posteriormente, os valores de **β**1 obtidos foram aplicados à seguinte fórmula, em vista da identificação das taxas de variação:

Rate = [-1 + e^**β**1^] X 100

The confidence intervals (CI) of the percent change rates were calculated based on the maximum and minimum **β** values using the following formula:

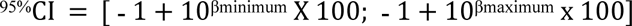

The null hypothesis (H0) was that the trend is stationary, i.e. there is no significant difference between the percent change rate and zero, adopting a 95% confidence level. Depending on the percent change rate, the trend may either be increasing (positive value), decreasing (negative value), or stationary, when the null hypothesis is accepted. Data processing and analysis was performed using SPSS and R.

## Results

Table 2 shows that the annual number of live births fell by 8.1% over the study period. The proportion of teenage mothers decreased over the period, while the percentage of mothers aged 35 years and over, mothers with a higher level of education, and multiple pregnancies increased. Deliveries were predominantly caesarean sections, with rates remaining constantly above 50%, although the proportion of this type of delivery fell slightly over the study period. The proportion of pre-labor caesarean sections was high, with rates reaching almost 70% throughout the time series.

**Table 2.**
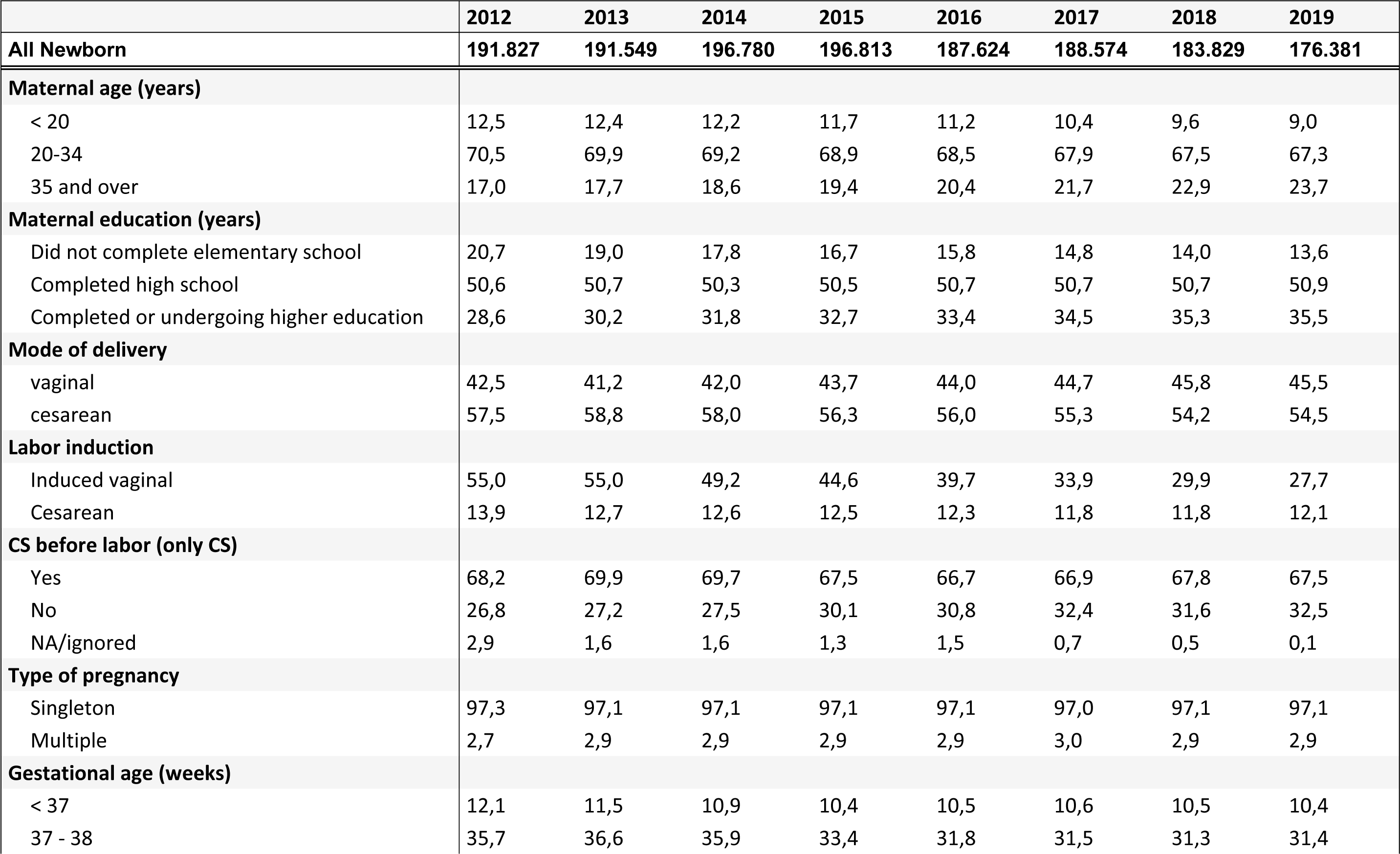
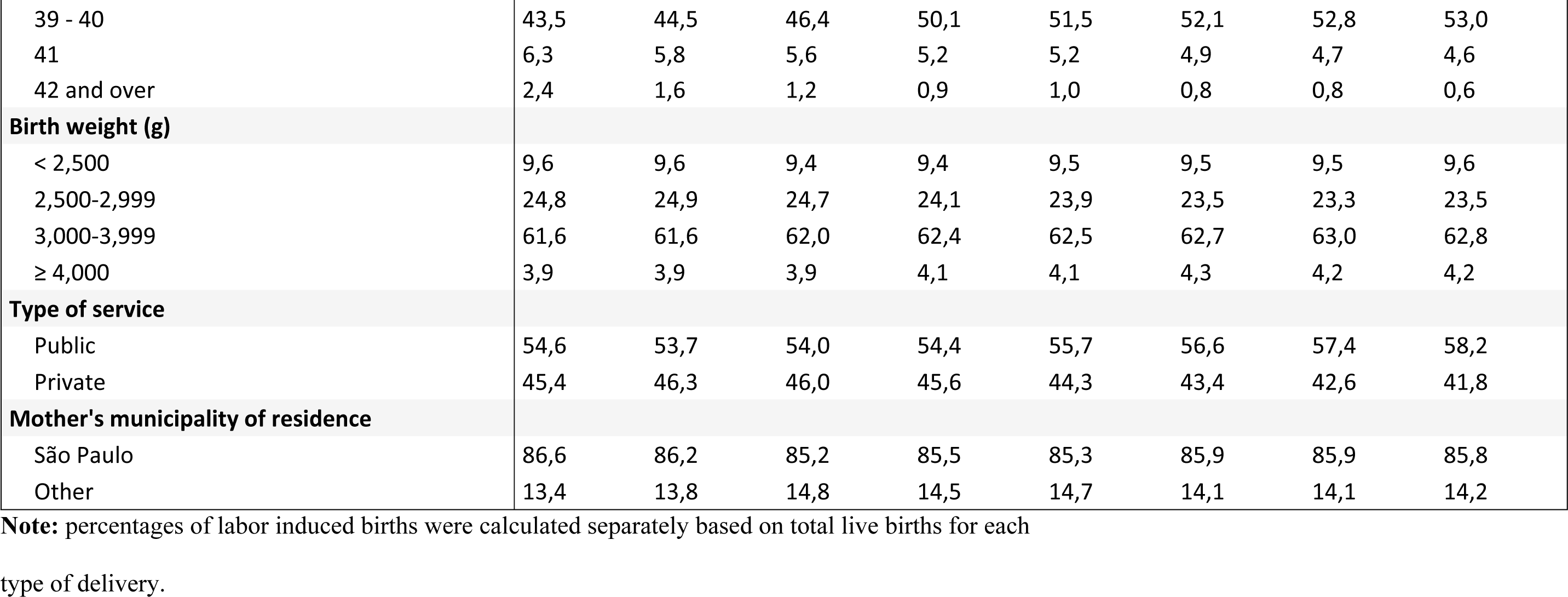
Annual number of live births and proportions according to maternal sociodemographic and birth characteristics. City of São Paulo, Brazil, 2012-2019.

The proportion of induced labor vaginal births fell over the period, from 55.0% in 2012 to 27.7% in 2019. With regard to length of gestation, the rate of full-term births (39-40 weeks) increased from 43.5% in 2012 to 53.0% in 2019, while the proportion of births at other gestational ages decreased. The rate of preterm births fell from 12.1% in 2012 to 10.4% in 2015 and remained relatively stable thereafter (Table 2).

Figure 2 shows that the proportion of live births between 30 and 35 weeks remained stable over the study period, while the proportion of births between 36 and 38 weeks fell from 2015, when the percentage of births between 39 and 40 weeks increased.

Figure 3 shows that the most common GA for both spontaneous and induced labor vaginal births was 39 weeks throughout the period. There was an increase in the proportion of spontaneous vaginal births at 39 and 40 weeks over the study period. The proportion of induced labor vaginal births at 39 weeks increased up to 2015, while the proportion of this type of birth at 40 weeks increased over the whole period at a proportionately higher rate than spontaneous vaginal births.

**Figure 3.**
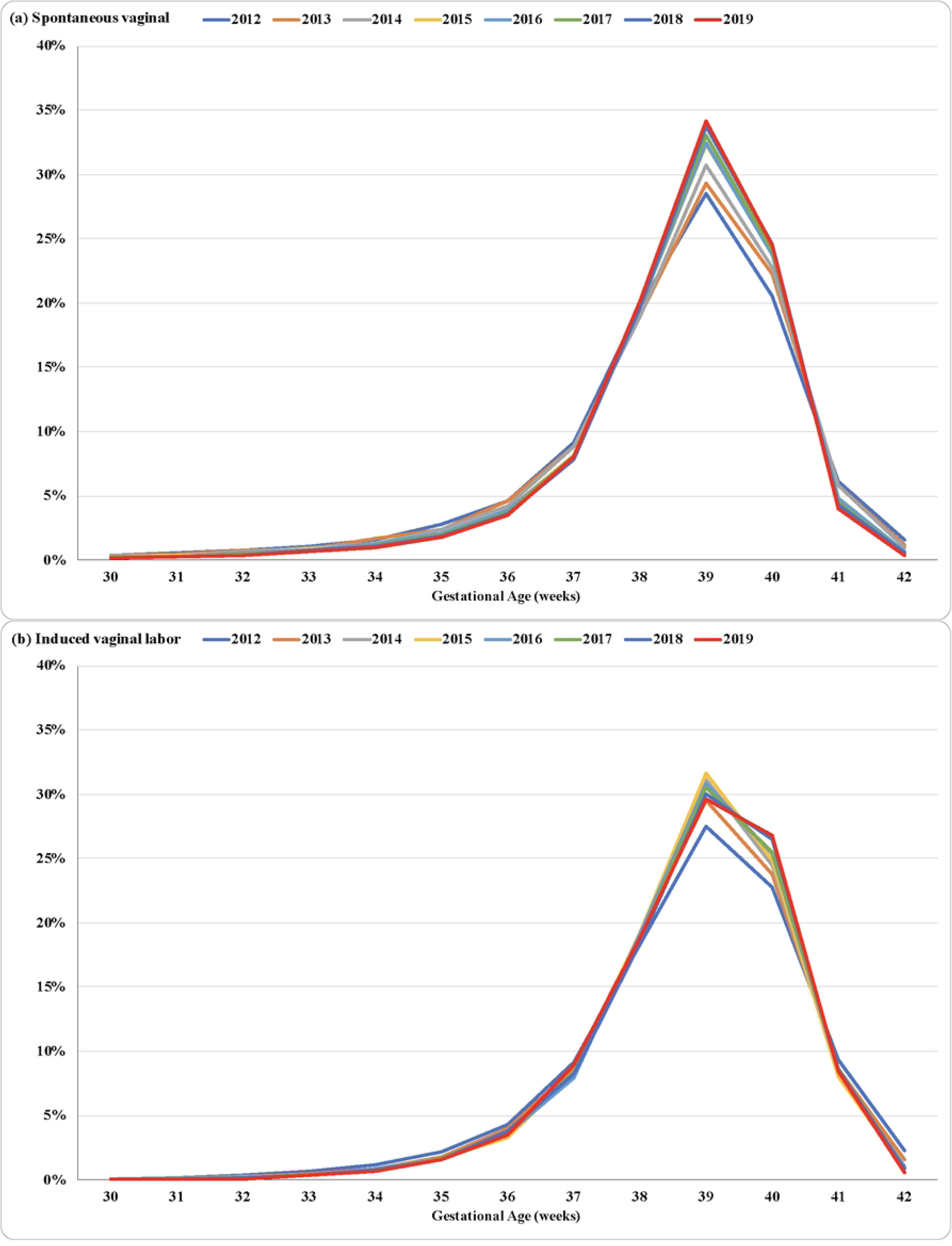
Distribution of spontaneous (a) and induced labor (b) vaginal births by gestational age. City of São Paulo, 2012-2019.

There was an increase in the proportion of intrapartum caesarean sections at 39 and 40 weeks up to 2015, with rates levelling off in the second four-year period, especially for births at 40 weeks. The most common gestational age for pre-labor caesarean sections was 38 weeks up to 2015, shifting to 39 weeks between 2016 and 2019, with rates remaining above 30% (Fig 4).

**Figure 4.**
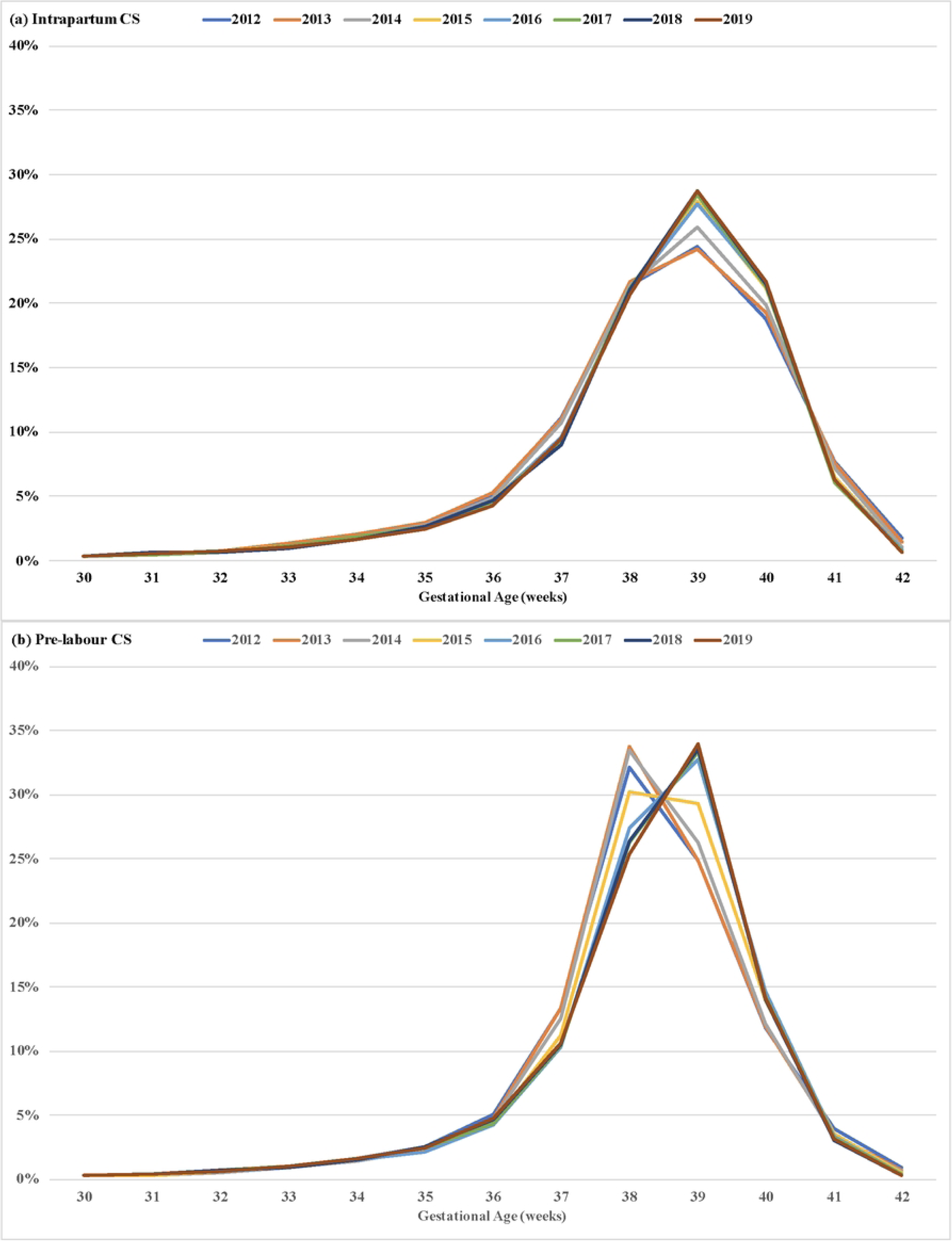
Distribution of intrapartum (a) and pre-labor (b) caesarean section births by gestational age. City of São Paulo, 2012-2019.

There was an increase in the proportion of births at 39 and 40 weeks in public hospitals and normal birth centers over the study period, while in private facilities there was a reduction in the proportion of births at 38 weeks from 2015 and increase in the percentage of births at 39 weeks (Fig 5).

**Figure 5.**
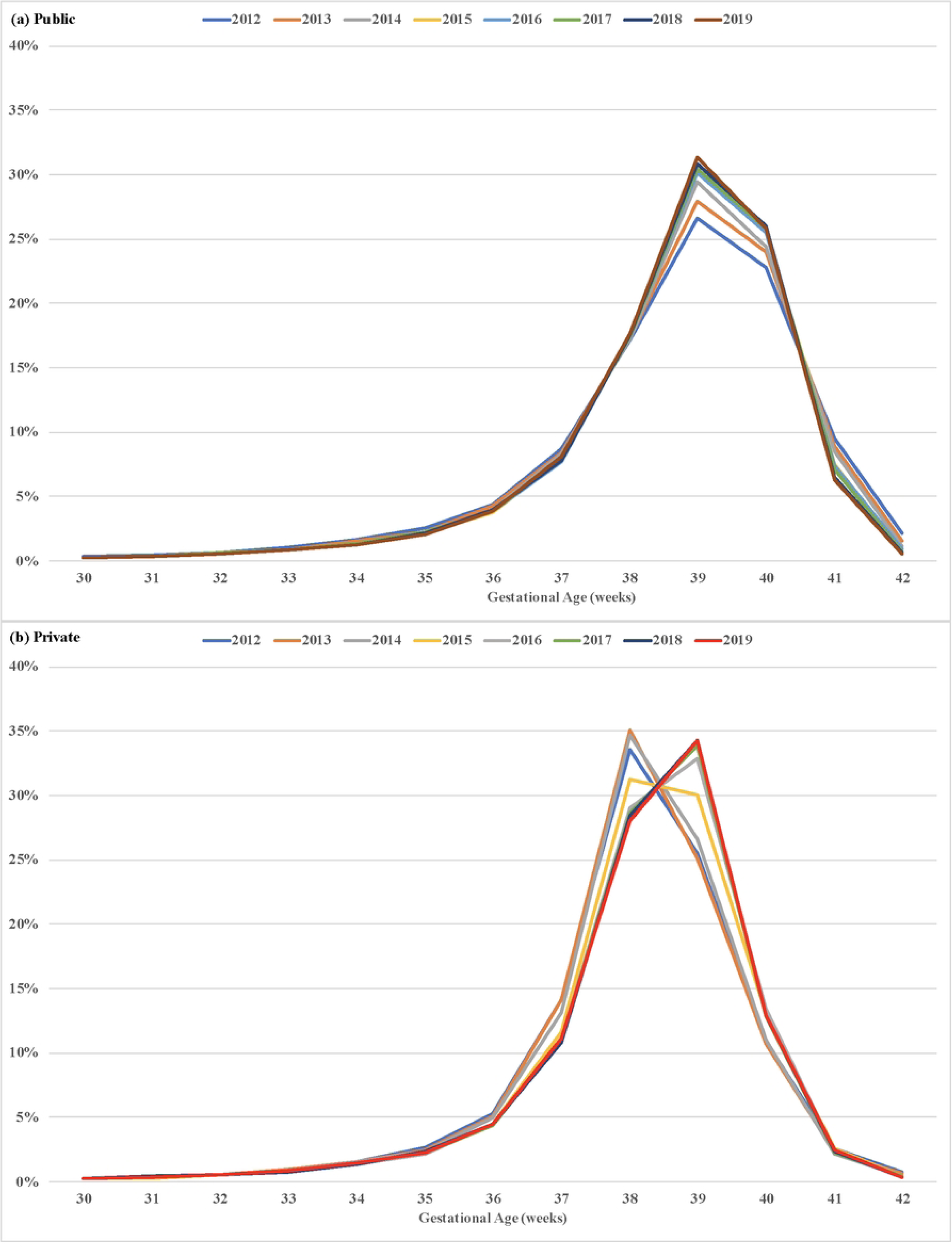
Distribution of live births in public (a) and private (b) hospitals by gestational age. City of São Paulo, Brazil, 2012-2019.

The average annual increase in the proportion of births at 39 and 40 weeks was 7.9% and 5.7%, respectively, while the proportion of births at other gestational ages showed a statistically significant reduction over the study period. These reductions were more pronounced in the first four-year period (2012-2015). There was also a substantial decline in the proportion of births at less than 37 weeks gestation over the study period. This reduction was also more pronounced in the period 2012-2015 (Table 3).

**Table 3.** Average annual percent change in proportion of live births by gestational age. City of São Paulo, 2012-2019.

| GA | 2012 to 2015 |  | 2016 to 2019 |  | 2012 to 2019 |  |
| --- | --- | --- | --- | --- | --- | --- |
|  | rate (%) | 95% CI | rate (%) | 95% CI | rate (%) | 95% CI |
| < 34 | -9,2 * | (-13.1;-5.1) | -2,7 * | (-2.7;-2.7) | -3,4 * | (-6.0;-0.7) |
| 34 | -3,4 | (-26.3;26.7) | -5,4 * | (-5.4;-5.4) | -4,3 * | (-8.4;0.0) |
| 35 | -11,1 * | (-17.4;-4.3) | -2,1 | (-11.0;7.7) | -5,4 * | (-8.9;-1.7) |
| **36** | **-10,7 \*** | (-15.1;-6.0) | **1,6 \*** | (1.6;1.6) | **-4,7 \*** | (-8.3;-1.0) |
| **37** | **-10,7 \*** | (-17.6;-3.2) | **-0,2 \*** | (-0.2;-0.2) | **-6,2 \*** | (-10.7;-1.5) |
| **38** | **-2,5** | (-11.4;7.2) | **-2,1 \*** | (-2.8;-1.3) | **-4,9 \*** | (-7.5;-2.3) |
| **39** | **12,2 \*** | (6.6;18.1) | **2,8 \*** | (2.1;3.6) | **7,9 \*** | (4.4;11.5) |
| **40** | **6,4 \*** | (6.4;6.4) | **1,6 \*** | (1.6;1.6) | **5,7 \*** | (2.8;8.6) |
| **41** | **-12,7 \*** | (-14.6;-10.8) | **-8,4 \*** | (-10.4;-6.3) | **-9,2 \*** | (-10.7;-7.7) |
| **42 and over** | **-52,8 \*** | (-55.2;-50.3) | **-26,2 \*** | (-26.2;-26.2) | **-33,6 \*** | (-43.0;-22.7) |

\*p < 0.05 rejects the null hypothesis.

Table 4 shows that there was a positive annual percent change in the proportion of vaginal births at 38 to 40 weeks over the study period. For births at 37 weeks, the annual percent change over the study period was negative (−7.3%) for caesarean births and positive for vaginal births (+1.2%).

**Table 4.**
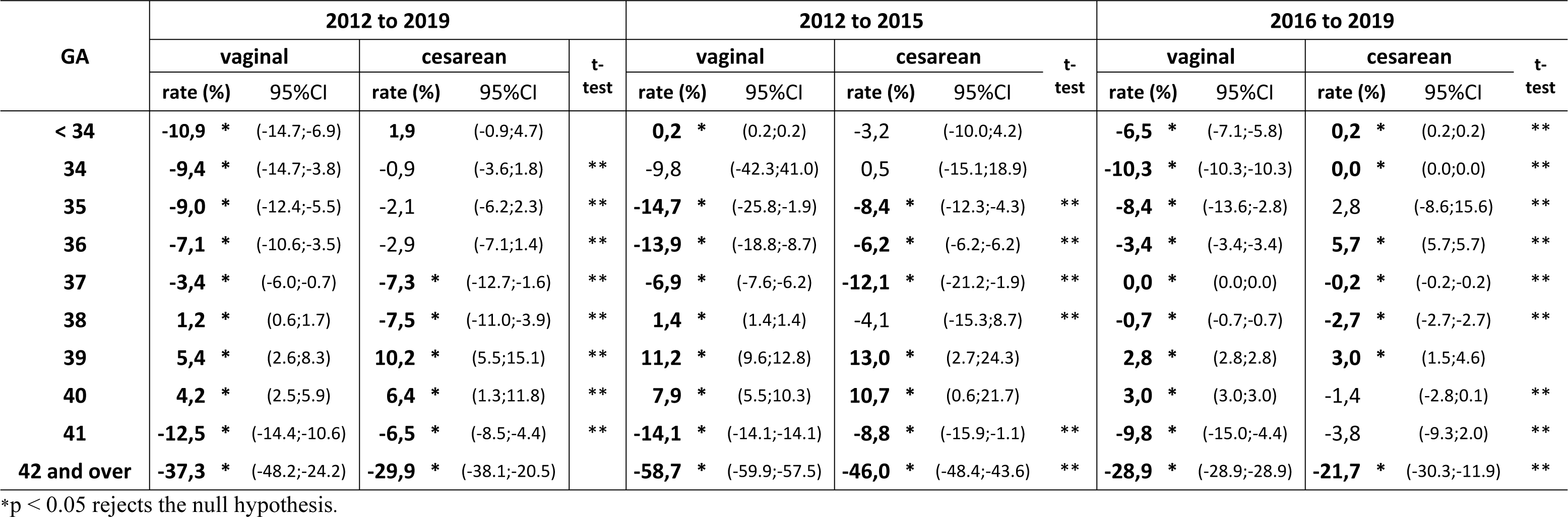
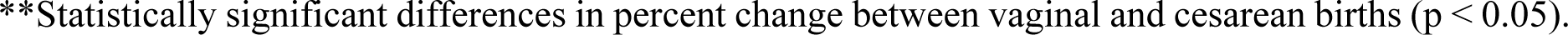
Annual percent change in the proportion of live births according to gestational age by type of delivery. City of São Paulo, 2012-2019.

In the period 2012-2015, the annual percent change in the proportion of caesarean births at 37 weeks was −12.1%, compared to −6.9% for vaginal births. This difference was statistically significant. The proportion of preterm births decreased across all gestational ages under 37 weeks. This reduction was greater in vaginal births.

Table 5 shows that there was a positive annual percent change in the proportion of births at 39 and 40 weeks throughout the study period for births in both public and private hospitals. This change was more pronounced in the period 2012-2019 and for births in private facilities. The proportion of births at 37 weeks decreased over the period 2012-2015 in both public and private hospitals (−5.4% versus 14.1%), while the proportion of births at 39 weeks increased (+11.9% versus +13.5%). The findings also show that there was a reduction in the proportion of late preterm births in both public and private hospitals (Table 5).

**Table 5.**
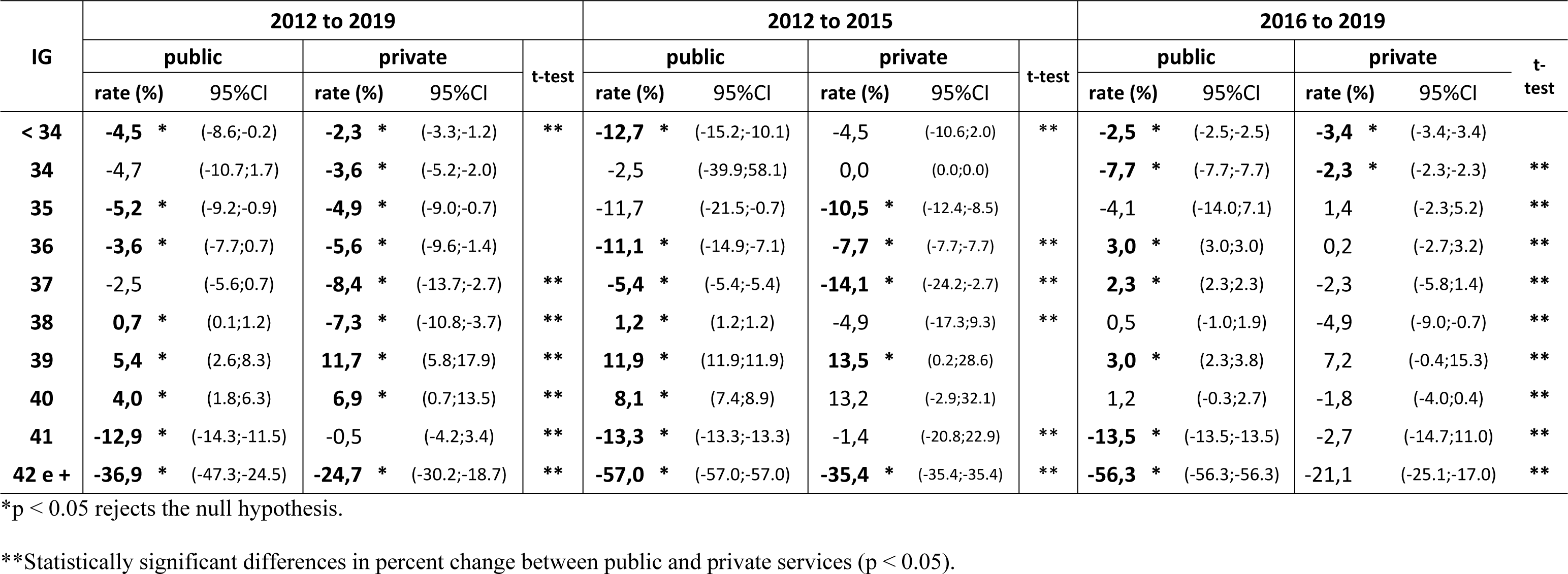
Annual percent change in the proportion of live births according to gestational age by system (public or private). City of São Paulo, 2012-2019.

Table 6 shows that there was a significant positive annual percent change in the proportion of births at 39 and 40 weeks across all age groups and levels of education. Rates were higher among mothers aged 35 years and over and who had completed more than 12 years of education.

**Table 6.** Annual percent change in the proportion of live births according to gestational age by maternal age and education level. City of São Paulo, 2012-2019.

| GA | maternal age (years) |  |  |  |  |  |
| --- | --- | --- | --- | --- | --- | --- |
|  | < 20 |  | 20 to 34 |  | 35 and over |  |
|  | rate (%) | 95%CI | rate (%) | 95%CI | rate (%) | 95%CI |
| **< 34** | **-4,5 \*** | (-8.6;-0.2) | **-3,6 \*** | (-5.7;-1.5) | **-2,3** | (-5.9;1.5) |
| **34** | **-5,8** | (-13.2;2.2) | **-5,2 \*** | (-8.2;-2.0) | **-3,2** | (-8.8;2.8) |
| **35** | **-7,3 \*** | (-9.3;-5.3) | **-5,2 \*** | (-8.7;-1.5) | **-5,4** | (-10.4;-0.0) |
| **36** | **-4,7 \*** | (-6.8;-2.6) | **-5,8 \*** | (-9.3;-2.2) | **-4,7** | (-9.8;0.6) |
| **37** | **-5,4 \*** | (-6.4;-4.3) | **-7,3 \*** | (-11.7;-2.7) | **-6,9 \*** | (-11.8;-1.7) |
| **38** | **0,7** | (-0.4;1.8) | **-6,2 \*** | (-8.8;-3.7) | **-5,6 \*** | (-9.1;-1.9) |
| **39** | **5,4 \*** | (3.2;7.8) | **8,1 \*** | (5.2;11.1) | **9,1 \*** | (3.9;14.6) |
| **40** | **4,5 \*** | (1.7;7.4) | **7,2 \*** | (4.3;10.1) | **7,9 \*** | (4.4;11.5) |
| **41** | **-12,3 \*** | (-15.1;-9.4) | **-7,5 \*** | (-9.0;-6.0) | **-5,2 \*** | (-7.7;-2.5) |
| **42 and over** | **-35,9 \*** | (-45.2;-24.9) | **-32,1 \*** | (-42.3;-20.0) | **-32,5 \*** | (-40.8;-23.1) |

| GA | education (years) |  |  |  |  |  |
| --- | --- | --- | --- | --- | --- | --- |
|  | No education/up to 8 |  | 9 to 11 |  | 12 and over |  |
|  | rate (%) | 95%CI | rate (%) | 95%CI | rate (%) | 95%CI |
| **< 34** | **-3,8** | (-8.9;1.5) | **-3,4** | (-7.0;0.4) | **-1,8** | (-3.9;0.3) |
| **34** | **-6,0** | (-13.4;2.0) | **-3,8** | (-7.9;0.4) | **-3,8 \*** | (-5.4;-2.3) |
| **35** | **-3,6** | (-9.2;2.3) | **-4,1 \*** | (-7.6;-0.3) | **-6,9 \*** | (-10.4;-3.3) |
| **36** | **-2,7** | (-7.9;2.7) | **-4,9 \*** | (-8.5;-1.2) | **-6,0 \*** | (-10.0;-1.8) |
| **37** | **-2,3** | (-5.9;1.5) | **-6,0 \*** | (-9.5;-2.4) | **-9,6 \*** | (-15.4;-3.5) |
| **38** | **0,7 \*** | (0.7;0.7) | **-4,7 \*** | (-6.3;-3.2) | **-9,4 \*** | (-13.3;-5.4) |
| **39** | **5,7 \*** | (2.8;8.6) | **7,2 \*** | (4.3;10.1) | **10,4 \*** | (4.6;16.6) |
| **40** | **4,0 \*** | (0.1;8.0) | **5,7 \*** | (3.4;8.0) | **14,3 \*** | (7.1;22.0) |
| **41** | **-14,1 \*** | (-15.0;-13.2) | **-9,6 \*** | (-11.1;-8.1) | **9,6 \*** | (5.5;13.9) |
| **42 and over** | **-36,9 \*** | (-46.1;-26.1) | **-33,3 \*** | (-42.7;-22.3) | **-18,5 \*** | (-26.1;-10.1) |

\*p < 0.05 rejects the null hypothesis.

## Discussion

Our findings show that there was a decrease in the proportion of preterm (< 37 weeks) and early term (37-38 weeks) births between 2012 and 2019 and an increase in the proportion of births at 39 and 40 weeks from 2015. The caesarean section rate declined over the study period. However, this method remained the most common type of delivery, with rates being higher in private hospitals. The increase in the proportion of births at 39 and 40 weeks was higher for births by caesarean section.

The results also show that there was a reduction in the proportion of labor induced births over the study period. This may be partially explained by the modification to the CLB in 2011. Data quality has improved over time, especially in the city of São Paulo, where health professionals have received training on how to complete the form and the importance of differentiating between induced and natural labor [17].

International studies have reported a left shift in GA at birth [1,7] and increase in rates of late preterm and early term births related to a rise in obstetric interventions. Studies in Brazil [6,13,18], including São Paulo [5,15], have shown that scheduled caesarean sections, particularly in private hospitals, and induced labor early term births have contributed to shortening the length of gestation and a left shift in GA at birth. In contrast, the present study observed a right shift in the GA curve from 2015, with an increase in the proportion of births at 39 and 40 weeks and reduction in preterm and early term births. Our results also show that these changes were more pronounced for births in private hospitals. This trend may be partially attributed to a decline in obstetric interventions – such as labor induction and elective caesarean sections – particularly before 39 weeks of gestation. It is worth highlighting that the rate of increase in the proportion of births at 39 weeks was higher than the rate of fall in caesarean sections between 2012 and 2019.

It is reasonable to assume that factors other than the implementation of women’s health policies in the 1980s may have influenced the findings of the present study. In 2006, the women’s organization *Rede Parto do Princípio* reported the abuse of caesarean delivery in the private health system to the Public Prosecutor’s Office (MPF, acronym in Portuguese) [19,20]. The MPF accepted the complaint and filed a public interest civil lawsuit seeking to force the private health sector regulator, the *Agência Nacional de Saúde Suplementar* (ANS), to regulate the quality of obstetric services provided by the private health system [21], where caesarean section rates are particularly high, reaching 100% in some maternity units. On 6 January 2015, the ANS issued Normative Resolution 368 [22], which sets out measures to ensure that private health insurance policy-holders have access to data on caesarean section rates by healthcare operator and health facility and making the use of partographs and maternity notes mandatory. In March 2015, the ANS also launched the “Adequate Birth Project” [20] which promotes a set of strategies aimed at improving childbirth support to reduce caesarean section rates. In March 2016, the Federal Medical Council issued Resolution 2144 recommending that elective caesarean sections should only be performed from 39 weeks of gestation to guarantee the safety of the baby [23].

The combination of these factors helped draw attention to the excessive use of the caesarean section in the private sector, empowering women to make informed birth choices. Our findings show that 2015 was an inflection point for caesarean section rates in SP, with an increase in the proportion of this type of birth at 39 weeks up to the end of the time series. This rise was more pronounced in private sector births.

Early caesarean sections without a medical reason can have adverse effects on short- and long-term maternal and infant health and well-being. At the end of pregnancy, every day counts and the more the birth is brought forward the higher the risk of infant mortality [24]. Thus, monitoring trends in GA at birth can help assess not only chances of survival but also future health. The reduction in preterm and early term births and increase in the proportion of births at later GAs is therefore good news. Nevertheless, induced labor and caesarean births remain the most common mode of delivery in SP, showing that non-evidenced based practices prevail in the country’s most populous city.

There is still a long way to go to improve intrapartum care in Brazil, which entails the uptake of national and international recommendations [25–27], including:

1. the use of evidence-based practices, promoting a change in culture, where childbirth is not seen primarily as a high-tech medical event.
2. the dissemination of an antenatal intrapartum care model involving interdisciplinary teams, including autonomous obstetric nurses and midwives for low-risk pregnancies and the active participation of women in their childbirth [28].
3. the strengthening of the country’s public health system, the *Sistema Único de Saúde* (SUS) or Unified Health System, and its principles and guidelines, including public participation and the dissemination of information on women’s rights.
4. the transparent monitoring of data and service and professional performance indicators.
5. the regulation of intrapartum care practices and price setting in the private sector to ensure good ethical standards.

It is hoped that the effective implementation of these recommendations will make birth safer and improve women’s satisfaction with the childbirth experience, positively influencing long-term infant outcomes. This constitutes a broad health care agenda that merits priority attention if we want to promote the health of women and the next generations.

## Data Availability

The database is fully available for consultation at the Harvard Dataverse Repository: https://doi.org/10.7910/DVN/PP2VVF

## Notes

### Competing Interest Statement

The authors have declared no competing interest.

### Funding Statement

Yes

### Author Declarations

This study was approved by the Research Ethics Committee of the School of Public Health - University of São Paulo (CAAE: 98163018.2.0000.5421), on October 11, 2018. This study analyzed anonymized secondary data, therefore individual consent was not obtained.

